# Effect of calcium intake on iron absorption and hematologic status: A systematic review and dose-response meta-analysis of randomized trials and case-cross-over studies

**DOI:** 10.1101/2020.09.21.20198358

**Authors:** Ajibola Ibraheem Abioye, Taofik A Okuneye, Abdul-Majeed O Odesanya, Olufunmilola Adisa, Asanat I Abioye, Ayorinde I Soipe, Kamal A Ismail, JaeWon F Yang, Luther-King Fasehun, Moshood O. Omotayo

**Affiliations:** Department of Nutrition, Harvard T.H. Chan School of Public Health, Boston MA, USA; Department of Family Medicine, General Hospital, Odan, Lagos, Nigeria; St Helen’s & Knowsley Teaching Hospitals NHS Trust, UK; Emory University, Atlanta GA, USA; University of Rhode Island, Kingston RI, USA; Department of Emergency Medicine, Rhode Island Hospital, Providence RI, USA; Department of Hematology, Lagos State University College of Medicine, Lagos, Nigeria; Warren Alpert Medical School, Brown University, Providence RI, USA; Wellbeing Foundation Africa, Abuja, Nigeria; Centre for Global Health, Massachusetts General Hospital (MGH), Boston MA, USA; Department of Pediatrics, Harvard Medical School, Boston MA, USA

**Author notes:** Correspondence to: Moshood Omotayo, Centre for Global Health, Massachusetts General Hospital (MGH), Boston MA, USA. Authors’ names for Pubmed indexing: Abioye, Okuneye, Odesanya, Adisa, Abioye, Soipe, Ismail, Yang, Fasehun, Omotayo. Competing interests: All authors have completed the ICMJE uniform disclosure form at www.icmje.org/coi_disclosure.pdf and declare: no support from any organization for the submitted work; no financial relationships with any organizations that might have an interest in the submitted work in the previous three years; no other relationships or activities that could appear to have influenced the submitted work.

**Keywords:** Calcium and iron interaction, Calcium and preeclampsia, Maternal nutrition, Maternal Anemia, Meta-analysis

## Abstract

**Background:** The interaction between dietary (and supplementary) divalent ions has been a long-standing issue in human nutrition research. Developing optimal calcium and iron supplementation recommendation needs detailed knowledge of the potential trade-offs between: a) the clinical effects of concurrent intake on iron absorption and hematological indices, and b) the potentially negative effects of separated ingestion on adherence to either or both iron and calcium supplements. Human clinical studies have examined the effects of calcium intake on iron status, but there are no meta-analyses or recent reviews summarizing the findings.

**Objective:** We aimed to summarize the literature on the effect of calcium consumption from meals and supplements on iron indices in humans, and quantify the pooled effects.

**Design:** Peer-reviewed randomized and case-cross-over studies were included in this review.

**Result:** The negative effect of calcium intake was statistically significant in short-term iron absorption studies but the effect magnitude was low (weighted mean difference (WMD) = -5.57%, (95% CI: -7.09, -4.04)). The effect of calcium on iron status was mixed. There was a quadratic dose-response relationship between calcium intake and serum ferritin concentration. Higher daily calcium intake was associated with a modest reduction in serum ferritin concentration. There was, however, no reduction in hemoglobin concentration (WMD = 1.22g/L, 95% CI: 0.37, 2.07).

**Conclusion:** The existing body of studies is insufficient to make recommendations with high confidence due to heterogeneity in design, limitations of ferritin as an iron biomarker and lack of intake studies in pregnant women. Prescribing separation of prenatal calcium and iron supplements in free living individuals is unlikely to affect the anemia burden. There is a need for effectiveness trials comparing the effects of prescribing separated intake to concurrent intake, with functional end-points as primary outcomes, and adherence to each supplement as intermediate outcomes.

## Introduction

The interaction between dietary (and supplementary) divalent ions has been a long-standing issue in human nutrition research. Multiple studies have demonstrated that calcium inhibits iron absorption in short-term and single-meal studies (1-3). Studies that have measured the absorption ratio of iron in meals with different amounts of calcium have shown an inverse relationship(4). Two key mechanisms of action have been proposed for calcium-iron interaction(5). One potential mechanism is that luminal calcium leads to internalization of DMT1 receptors, limiting transfer of luminal iron into enterocytes. The other proposed mechanism is that calcium interferes with the transfer of iron across the enterocyte basolateral membrane. Recent reviews of inhibition mechanisms have provided some support for the first theory, but also suggested that homeostatic mechanisms compensate for the calcium-iron interaction, and the inhibitory effect is transient or at least not as clinically consequential as most short- term absorption studies would suggest(4).

Developing optimal calcium and iron supplementation recommendation requires detailed knowledge of the potential trade-offs between: a) the clinical effects of concurrent intake on iron absorption and hematological indices, and b) the potentially negative effects of separated ingestion on adherence to either or both iron and calcium supplements(6-8). While some studies have provided indication of the direction of these relationships, results are conflicting. Data from dietary and supplementation studies have been inconsistent, and it remains unclear whether there is a threshold dosage beyond which calcium exerts its inhibitory effects, the value of such threshold, and factors that might affect the threshold(9). Furthermore, longer-term studies that have examined the effect of calcium supplementation on haematological indices have reported conflicting results (10-14).

Prior narrative reviews have summarized existing studies of the interaction of dietary (and supplementary) calcium and iron(4, 15-17). While the narrative reviews were comprehensive, they often did not involve systematic and reproducible search strategy or meta-analysis. In addition, they did not include studies that have been published between 2010 and 2019, and we are unaware of any prior meta- analysis summarizing the clinical evidence on either of these issues. The objective of this study is to summarize human clinical studies that have examined the impact of calcium intake on iron status, identify factors moderating the effect and quantify the magnitude of the effect.

## Methods

We followed the Preferred Reporting Items for Systematic Reviews and Meta-analysis (PRISMA) guidelines in the design, analysis, and reporting of this study (**Supplement 1**)(18). We identified studies examining the impact of calcium intake on iron outcomes, including iron absorption ratio and hematological indices. Original peer-reviewed research articles published up to August 2020 in the following medical literature databases were identified and examined for inclusion in the review: PUBMED/Medline (U.S. National Library of Medicine) and EMBASE (Elsevier). The databases were searched using queries composed of MeSH terms (Medical Subject Headings), EmTree terms and keywords representing iron, calcium and absorption/bioavailability. Hand searching of references was also done – specifically by examining the references of relevant systematic reviews and included studies as well as the first 500 hits on Google Scholar. No restrictions by age, year of publication or language were implemented. The title and abstract of each study were screened and full-texts examined in duplicate **(Figure 1)**. A third author resolved discrepancies. Studies were excluded if they did not examine iron absorption or hematologic status, and did not examine calcium and iron intake, whether as supplements or in diet. If the diet differed by calcium intake and one or more other nutrient(s), the study was not included(19, 20).

**Figure 1.**
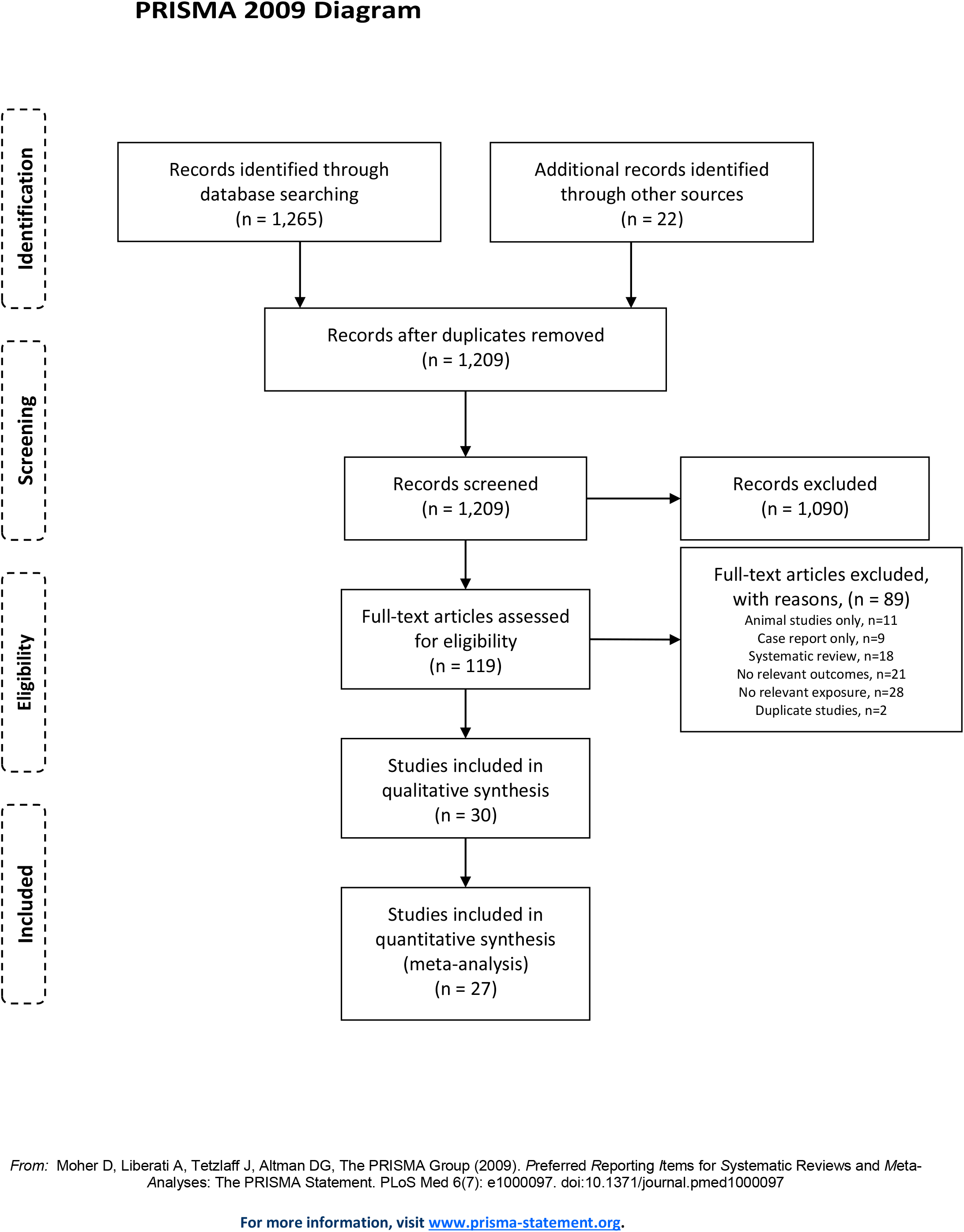
Flowchart of included studies

In terms of population, studies were not required to include only individuals who were healthy at baseline or to exclude individuals who were anemic or pregnant. Only randomized controlled trials and crossover studies were included, to facilitate causal interpretation of the effect of the interventions. The studies had to have compared different doses of calcium intake in supplements or meals or compared different sequences of calcium intake – keeping total daily intake constant.

### Data extraction

Data extraction from full-text articles was done using a comprehensive extraction sheet. Information on study design, population, intervention, covariates and findings were extracted. The median was extracted if the mean was missing. Intake of calcium was converted to mg/day. Included studies provided calcium in the test arm by altering the composition of meals, or providing calcium supplements. In some cases, numerical values of total daily calcium intake were not provided, and were therefore imputed by estimating the calcium content of meals provided using USDA reference(21). One study provided estimates from the same individuals when iron absorption studies were conducted with and without meals, and the estimates from studies without meal were included(2). If packed cell volume (PCV) was provided, missing hemoglobin data was estimated by dividing PCV by 3. Only two studies provided iron absorption estimates adjusted to ferritin 40µg/L, and the adjusted estimates were used (22, 23).

### Outcomes

The primary outcomes of interest were the iron absorption ratio (%) – total, heme and non-heme, serum ferritin (µg/L) and hemoglobin (g/L). While iron absorption ratio reflects short term effects, serum ferritin and hemoglobin reflect longer term effects. Regardless, all relevant outcomes reported in the included studies were reviewed. These include anemia (%), mucosal uptake of iron (% or mg), iron serosal transfer index, iron retention (mg), erythrocyte incorporation of iron (% of absorbed iron), zinc protoporphyrin (mmol/L), soluble transferrin receptor (µg/L), mean corpuscular volume (fL) and iron bioavailability (% of absorbed iron).

### Risk of bias assessment

We assessed the risk of bias of individual studies using the Cochrane risk of bias tool(24). For crossover trials, the Cochrane tool was modified as follows. Bias in randomization was described as high if the order of intervention was not randomized. In addition, bias due to carryover effects was assessed by the presence of absence of a washout period or a follow-up period non-interventional period (≥14 d) in the studies. Two investigators independently assessed the studies and disagreements were resolved by consensus with a third author. This ranking did not influence decisions concerning exclusion of studies or analytic approach.

### Statistical analysis

Included studies differed in the details of exposure assessment and outcome ascertainment. Random effects models, which explicitly model the between-study variation, were therefore selected *a priori* for the meta-analyses(25). Weighted mean differences for total, heme and non-heme iron absorption (%) and serum ferritin (µg/L) were obtained from pooled analysis of the highest daily calcium intake compared to the lowest daily calcium intake. For studies reporting the outcome of interest at multiple time points, the longest reported follow-up was included in the main analysis.

Heterogeneity was formally assessed with the I^2^ statistics, a measure of the total variability that is due to between-study variation. I^2^ was regarded as low if <50%, substantial if 50 – 90% and considerable is >90%, in accordance with the general guidelines for Cochrane reviews, and p-values for Q-statistic reported(24). Heterogeneity was further assessed using meta-regression approaches and analysis within subgroups defined by age (<18, 18 – 65 and ≥65 years), sex, nature of intervention (supplement or diet), and baseline iron status. The impact of an individual study on the WMD meta-analysis was evaluated by leaving one study out sequentially and obtaining pooled estimates. Publication bias was evaluated with funnel plots and Egger’s tests(26, 27).

Dose response meta-analysis of differences in means was conducted following the two-stage approach proposed by Crippa and Orsini, based on restricted maximum likelihood estimation method(28, 29). This approach makes no assumptions about the underlying shape of the association. Studies that did not report more than 2 categories from the same sets of individuals were not included in the dose-response meta-analysis (11, 23, 30). Included studies measured outcomes on similar, interpretable scales, and pooled difference estimates were therefore obtained on an absolute scale. Dose-response meta-analyses were conducted using a variety of regression approaches – restricted cubic splines, fractional polynomial and quadratic models, and the model with the lowest quantitative value of the Akaike Information Criteria (AIC) was selected as the final model. In the presence of nonlinearity, the selected model was presented in graphical form using predicted mean differences. Predicted mean differences for 1000, 1500 and 2000mg/d were obtained, in comparison to 500mg/d.

*P*-values are two sided and significance set at *p*<0.05. Statistical analyses were conducted using RStudio 1.0.153(*16*). Values presented in the text are means (±SD), means (95% CI), and means (±SE).

## Results

### Description of studies

We identified 30 papers from an initial set of 1287 titles and abstracts (**Figure 1**) reporting on the influence of consumption of calcium on iron absorption and hematologic indices. These were 12 randomized controlled trials(1, 10-12, 14, 22, 31-36) and 18 case-crossover studies(2, 23, 30, 36-48), including 1,623 and 592 participants respectively. The studies were conducted in Africa (14), Asia (10, 31, 36), Europe (1, 11, 12, 22, 23, 43), North America (2, 3, 13, 30, 34, 38, 39, 41, 44, 45, 47-49), and South America (32, 33, 42). Fourteen of the studies included women only (2, 3, 12-14, 22, 31, 32, 39, 41, 42, 45, 46, 48, 50, 51). Almost all studies explicitly excluded pregnant or lactating women or both (11, 12, 14, 22, 32, 33, 36, 42, 48) and none of the studies examined pregnant women alone. One study did not report the sex of participants (48). The interventions consisted of regular or low-calcium meals with or without calcium supplements (1-3, 11-14, 23, 30-33, 36, 39, 41, 42, 46, 48, 51), or high-calcium versus low-calcium meals (10, 22, 34, 38, 43-45, 47, 49, 52). The studies assessed iron absorption using radiolabeled iron (1, 2, 22, 32, 34, 36, 49) or gastrointestinal lavage and body scintillation procedures (30). Most of the studies were short-term and intervening over < 3 months (1, 3, 22, 30-32, 36, 38, 39, 41, 42, 44-49, 52), while others were longer-term and lasted 3 – 6 months (11, 33) or 1 – 4 years (12, 14, 51). Of these, two studies evaluated the outcome at multiple time points, allowing their inclusion in both short term and long-term analyses (11, 14). **Table 1** presents the characteristics of included studies.

**Table 1.**
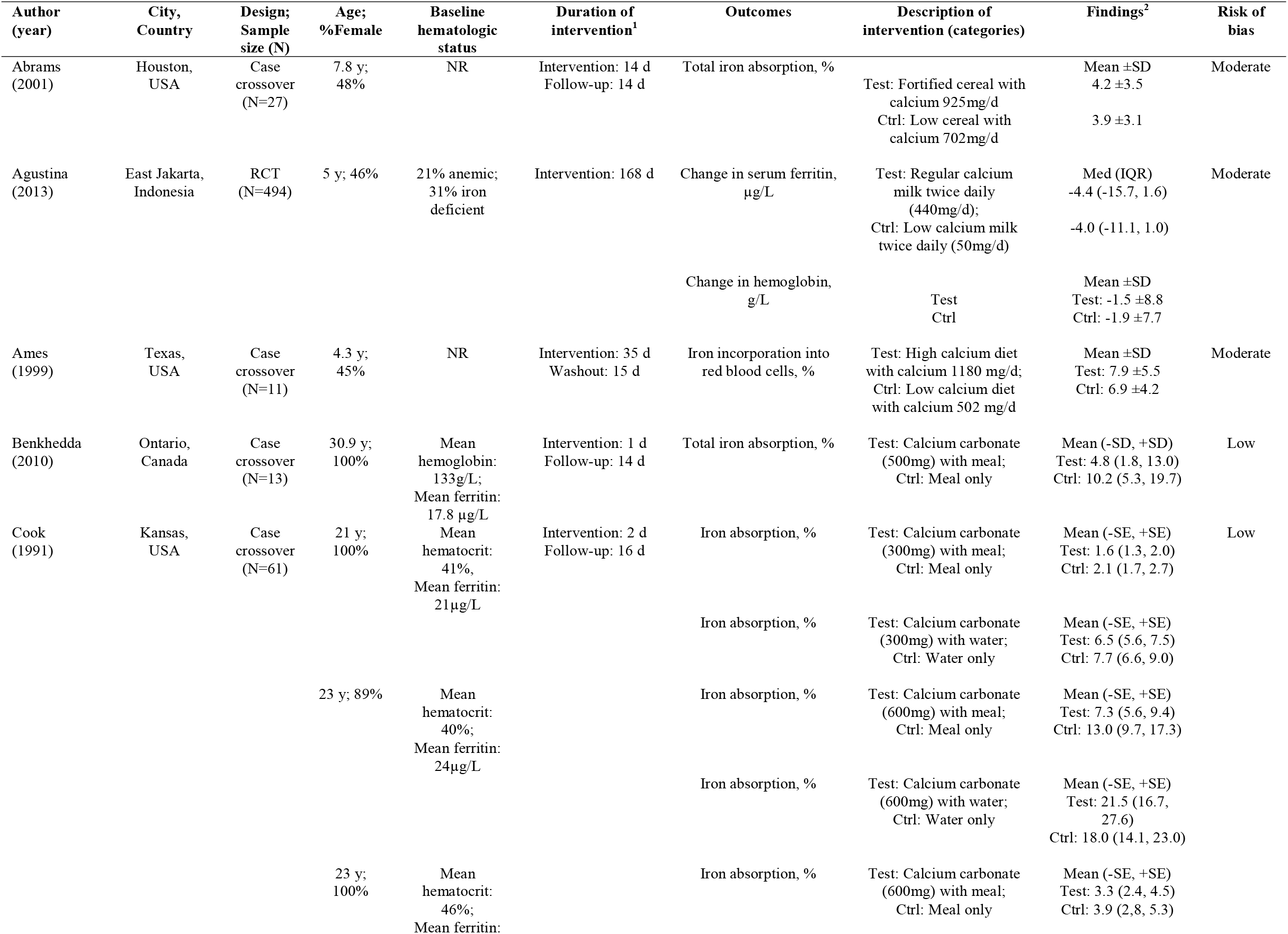
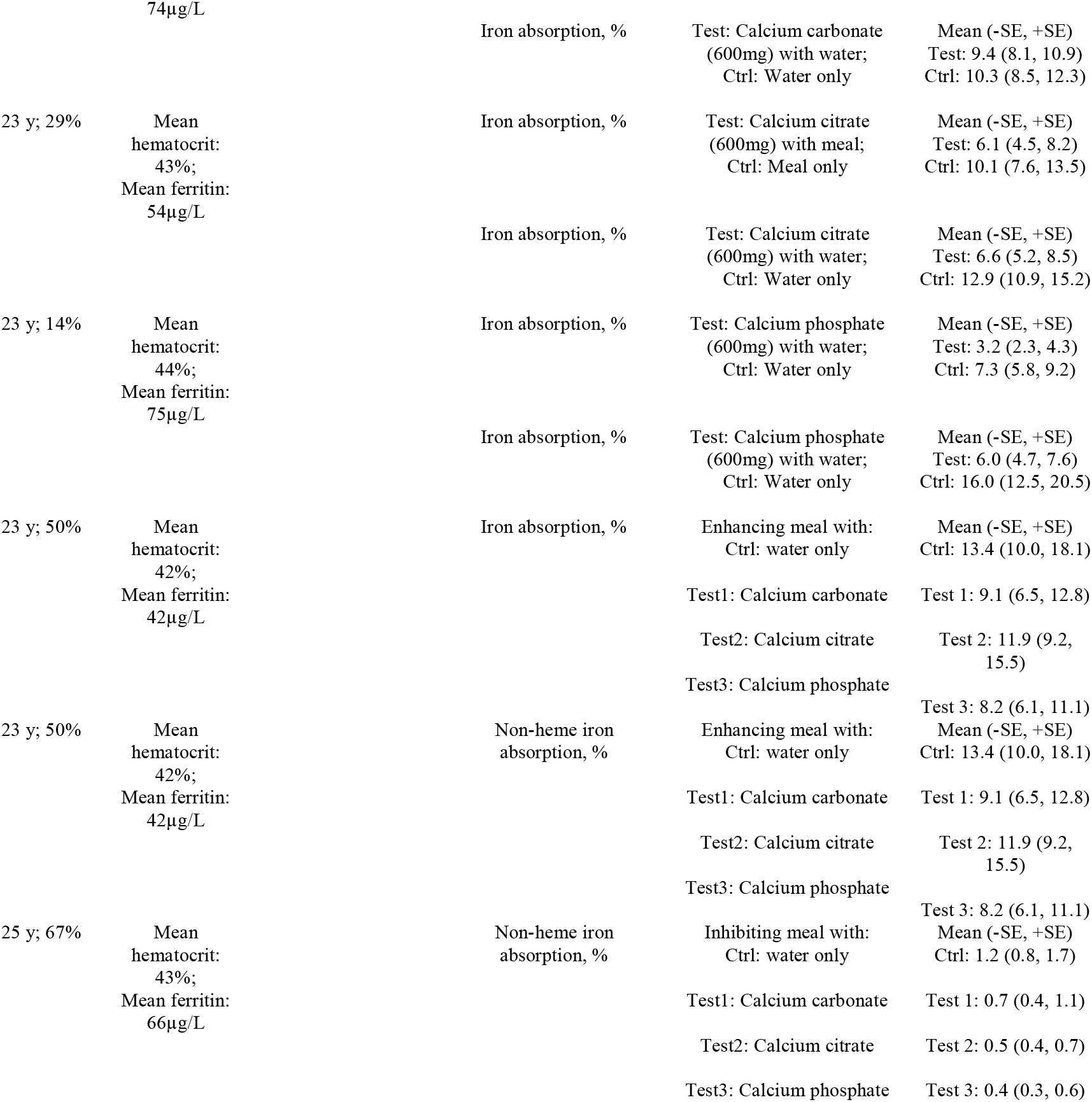
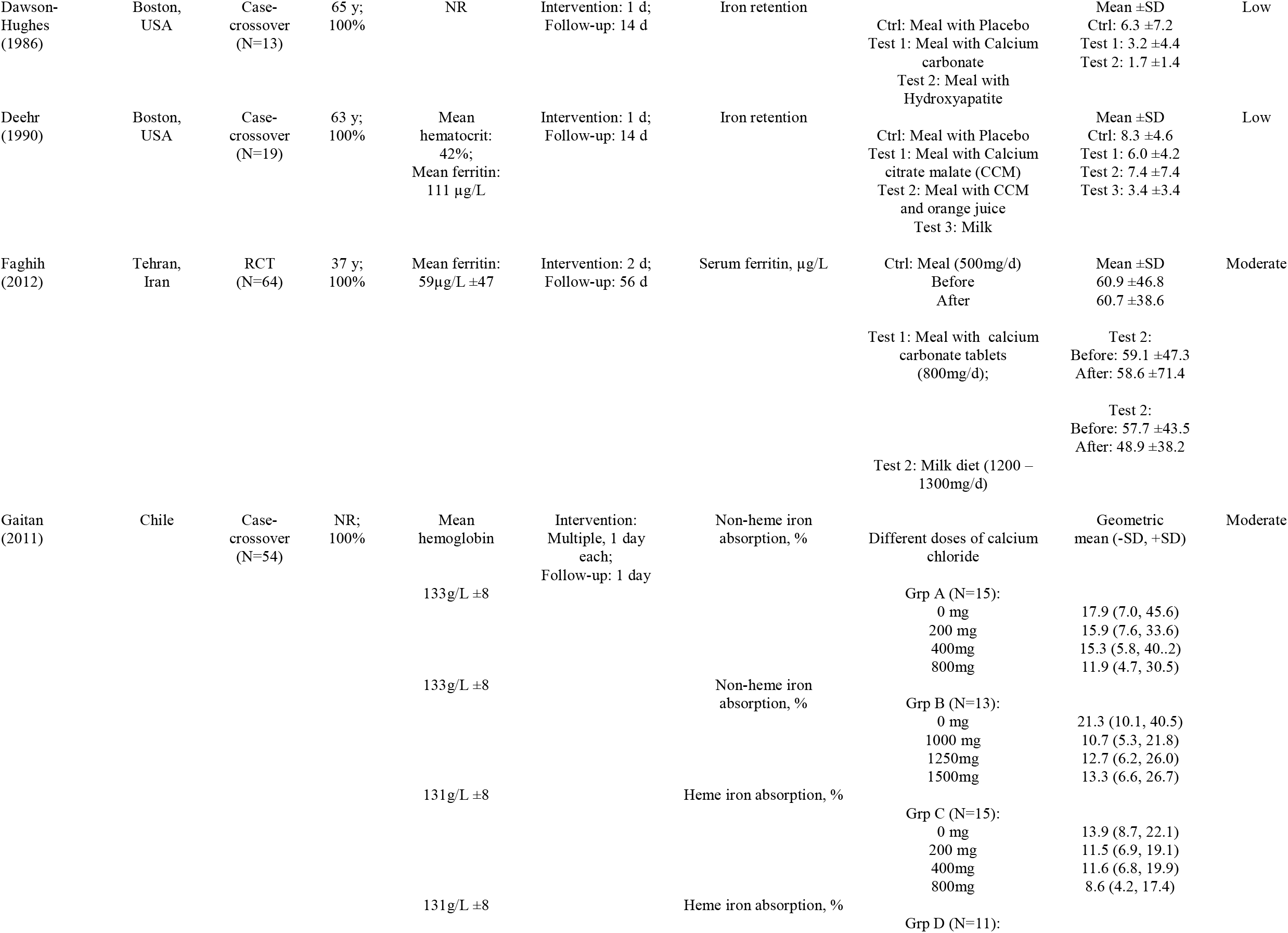
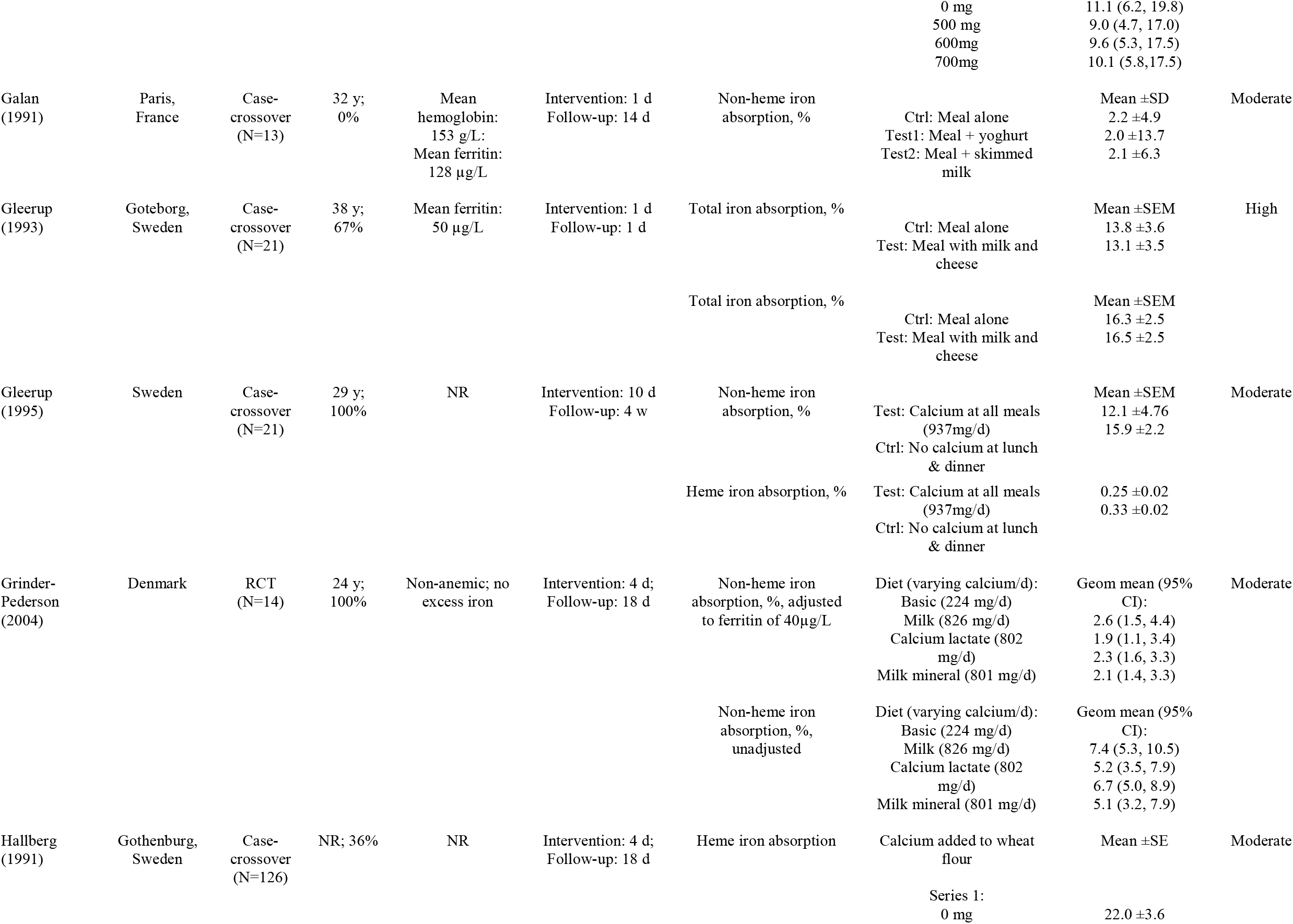
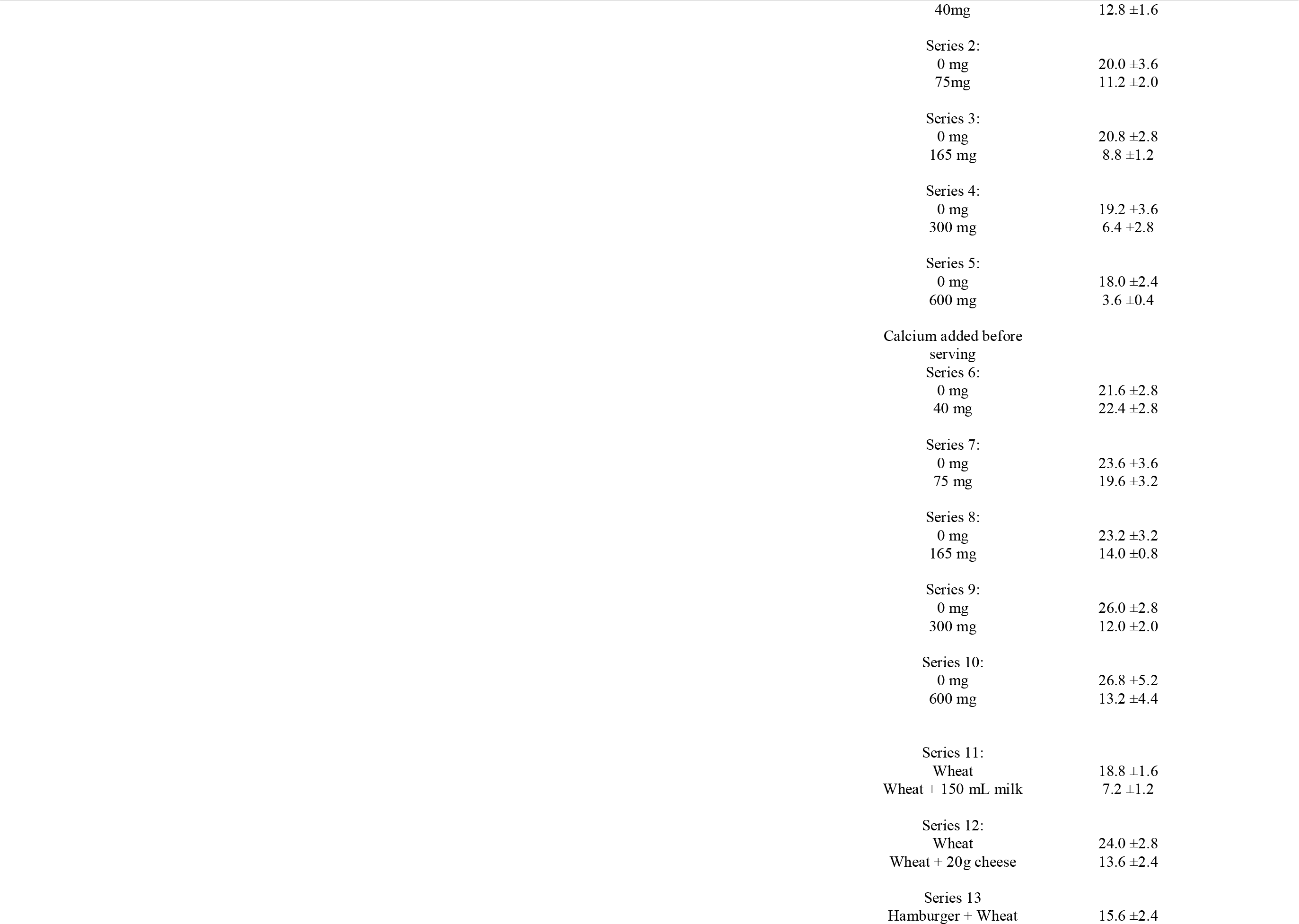
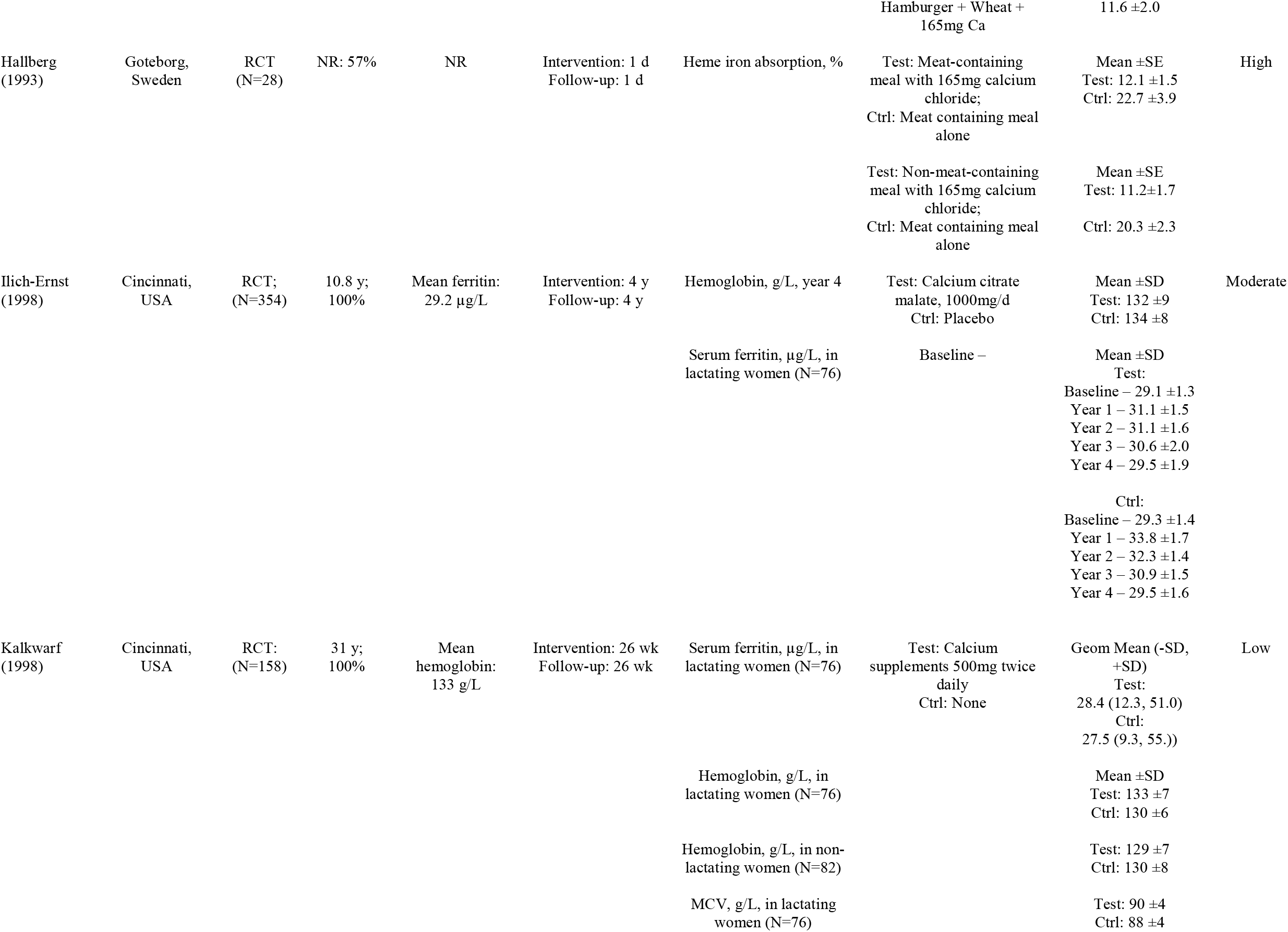
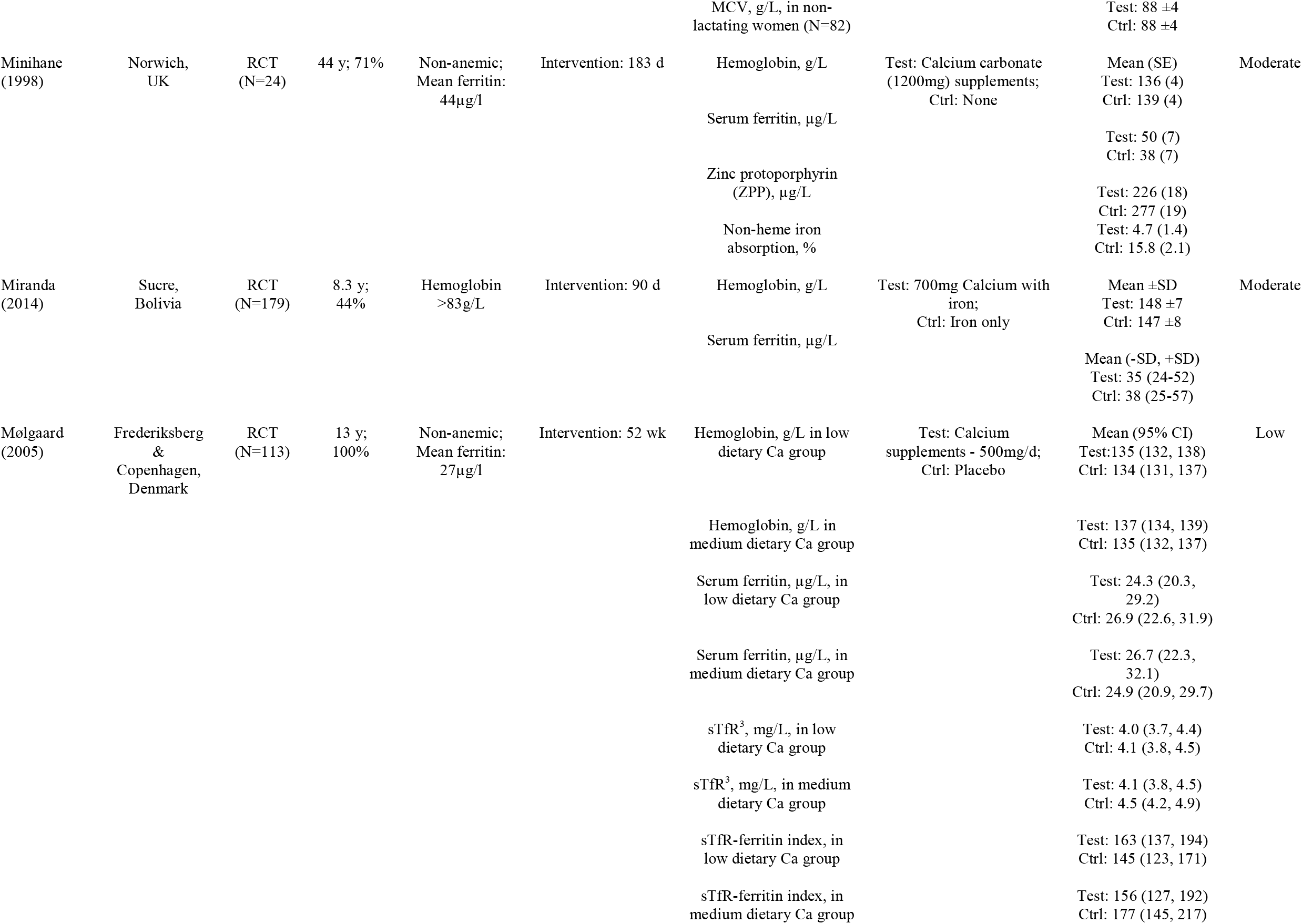
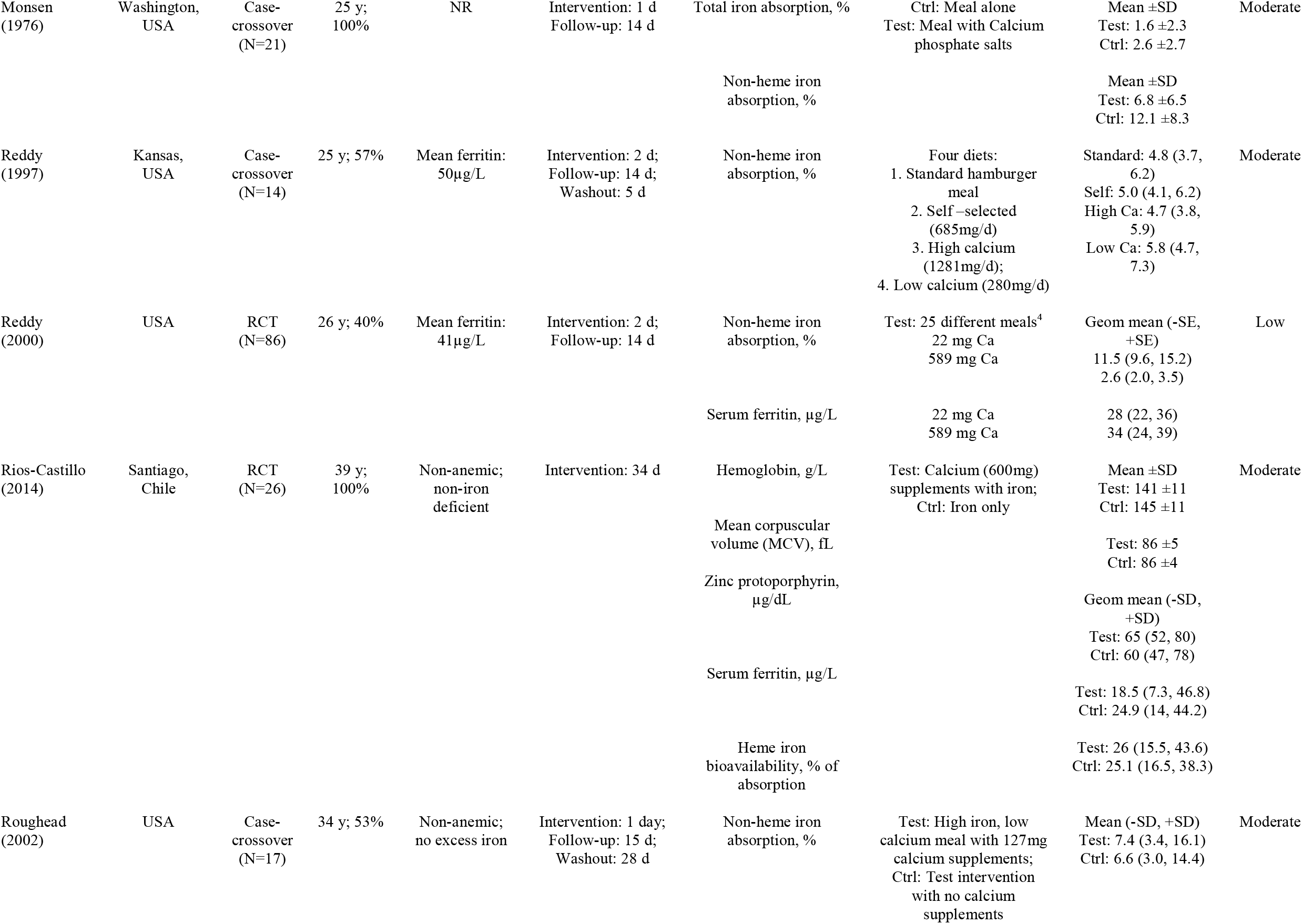
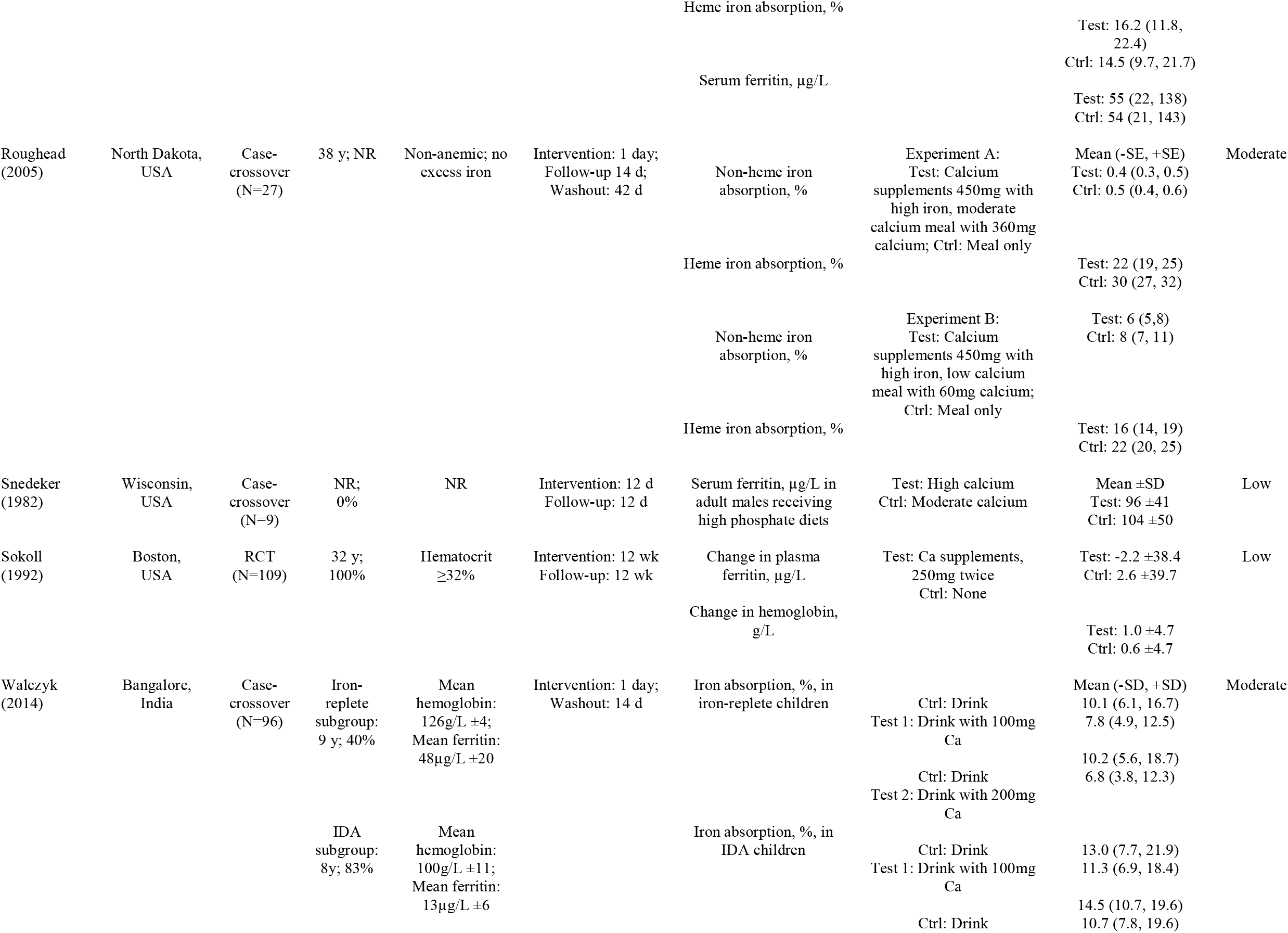
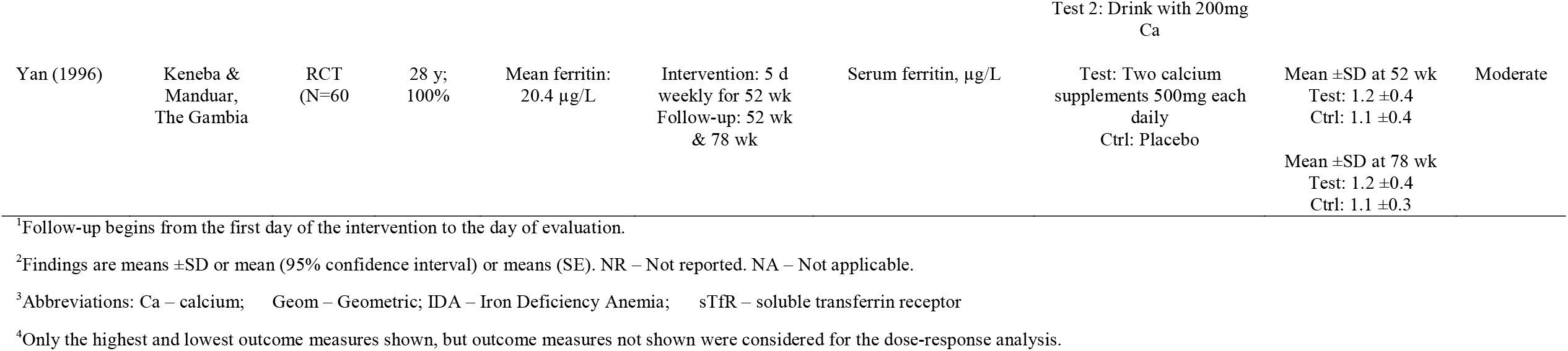
Characteristics of Included Studies.

Most of the included studies were of low risk of bias. The others were of moderate risk of bias (1, 10, 11, 14, 22, 30-33, 36, 42, 48, 51). Studies regarded as having a moderate risk of bias most often did not report (or perform) allocation concealment or blinding of participants and personnel.

### Total iron absorption

We pooled 24 estimates from 7 studies (1, 2, 11, 22, 23, 30, 34, 36, 39, 42, 43, 46-49) to obtain weighted mean differences comparing the influence of high calcium intake on iron absorption relative to low calcium intake, and found that calcium intake was associated with lower iron absorption (**Figure 2**. WMD = -5.91, 95% CI: -8.38, -3.44; I^2^=84%, *p*-heterogeneity<0.0001). Follow-up for the included studies was from 1 – 18 days. There was no evidence from influence analysis that the pooled estimate was dominated by any of the individual studies. Neither the funnel plot nor Egger’s mixed effects regression revealed any evidence of publication bias (p-value = 0.75; **Supplementary Figure 1**).

**Figure 2.**
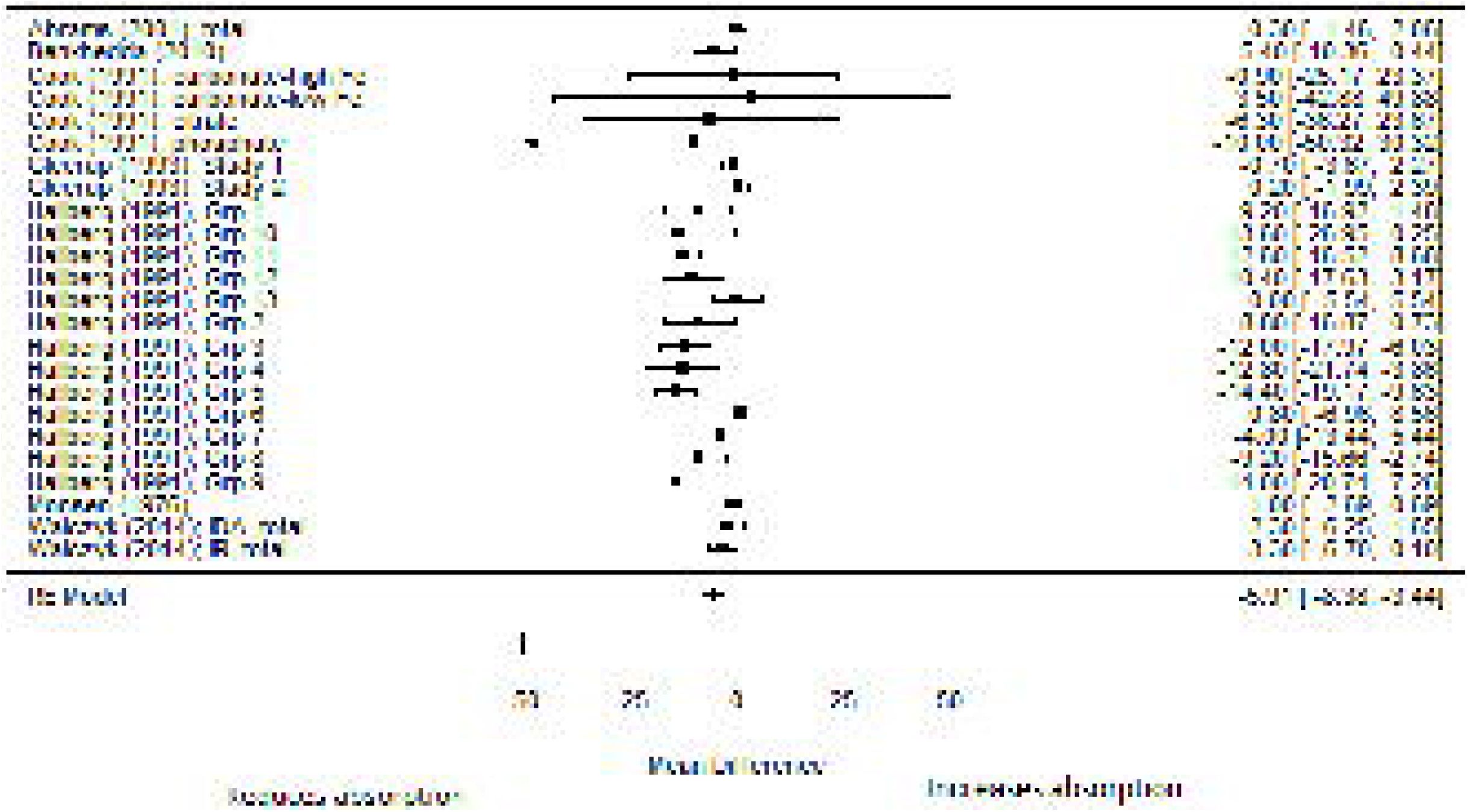
Forest plot for the relationship of calcium intake and total iron absorption (highest vs least calcium intake)

Heterogeneity was partly explained by the nature of the intervention (p<0.0001). The total iron absorption was -1.02 (95% CI: -3.77, 1.73) when intervention was based on meals alone, and differed by -7.90 (95% CI: -11.4, -4.3) when intervention was included supplements. Heterogeneity was not significantly explained by participant’s age (p-heterogeneity = 0.78), sex (p-heterogeneity=0.19), baseline serum ferritin (p-heterogeneity=0.13), hemoglobin concentration (p-heterogeneity=0.64), duration of follow-up (p-heterogeneity=0.19) or year of publication (p-heterogeneity=0.43).

We pooled 32 difference measures from 3 absorption studies(1, 2, 36) using quadratic model-based meta-analysis to evaluate the dose-response relationship of calcium intake and the daily iron absorption (**Figure 3)**. The dose-response association of calcium intake on total iron absorption was significantly non-linear – total iron absorption was poorer with higher calcium intake (*p-*value = 0.026).

**Figure 3.**
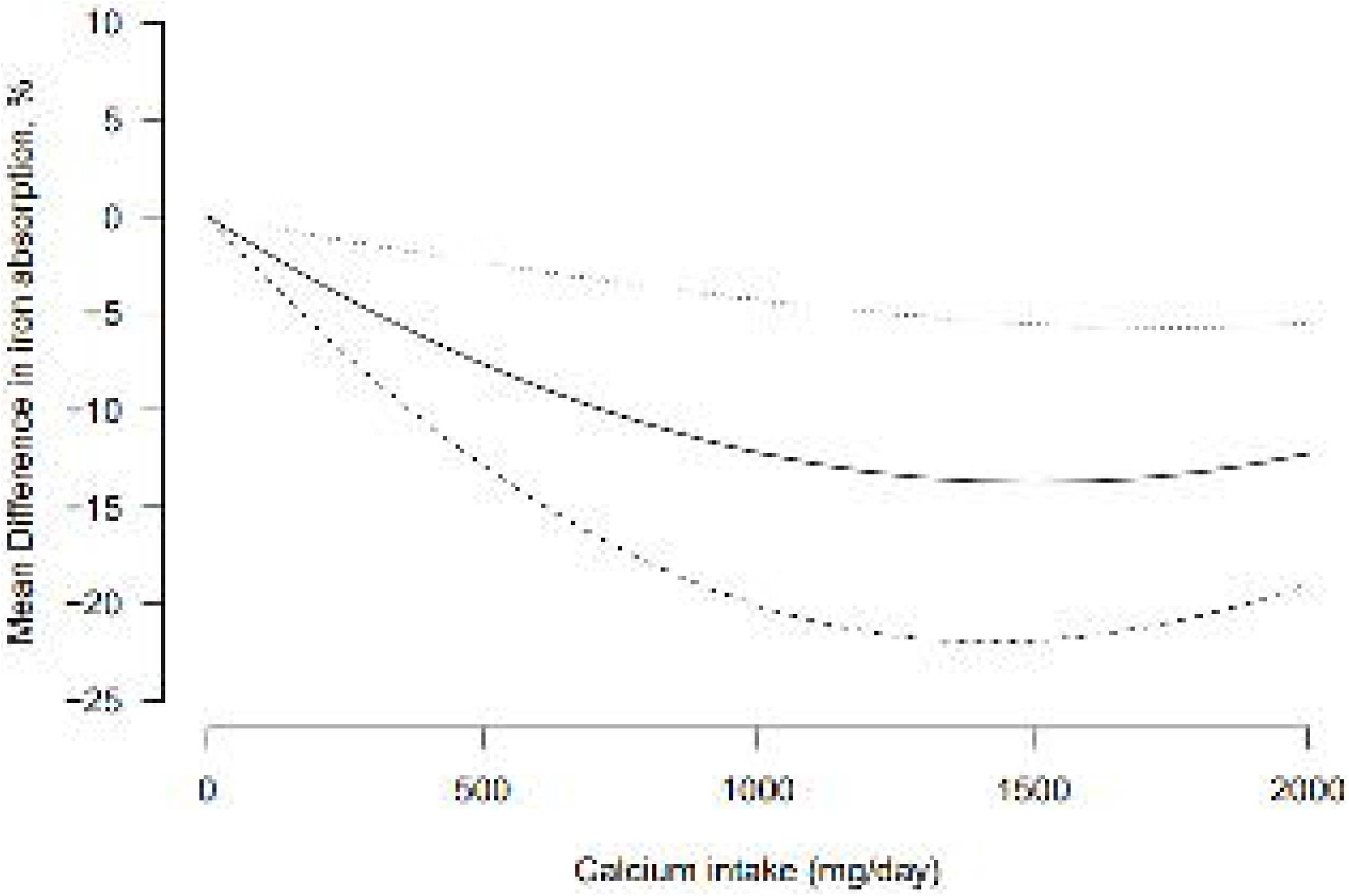
Dose-response relationship of calcium intake and total iron absorption

### Heme iron absorption

We pooled 7 estimates from 4 studies(1, 23, 42, 48) to obtain weighted mean differences comparing the influence of high calcium intake on heme iron absorption relative to low calcium intake, and found that calcium intake was associated with a slightly lower heme iron absorption (**Figure 4**, WMD = -4.84, 95% CI: -8.59, -1.09; I^2^=60%, *p*-heterogeneity=0.021). Follow-up of the included studies was from 1 – 18 days. All included studies had provided a supplement as the intervention. The influence of age, sex and baseline hematologic status on heterogeneity was not assessed due to missing covariate data. Visual inspection of the funnel plot suggests possible publication bias (**Supplementary figure 2)**, although there was no evidence from Egger’s test of small study effects (p-value=0.35).

**Figure 4.**
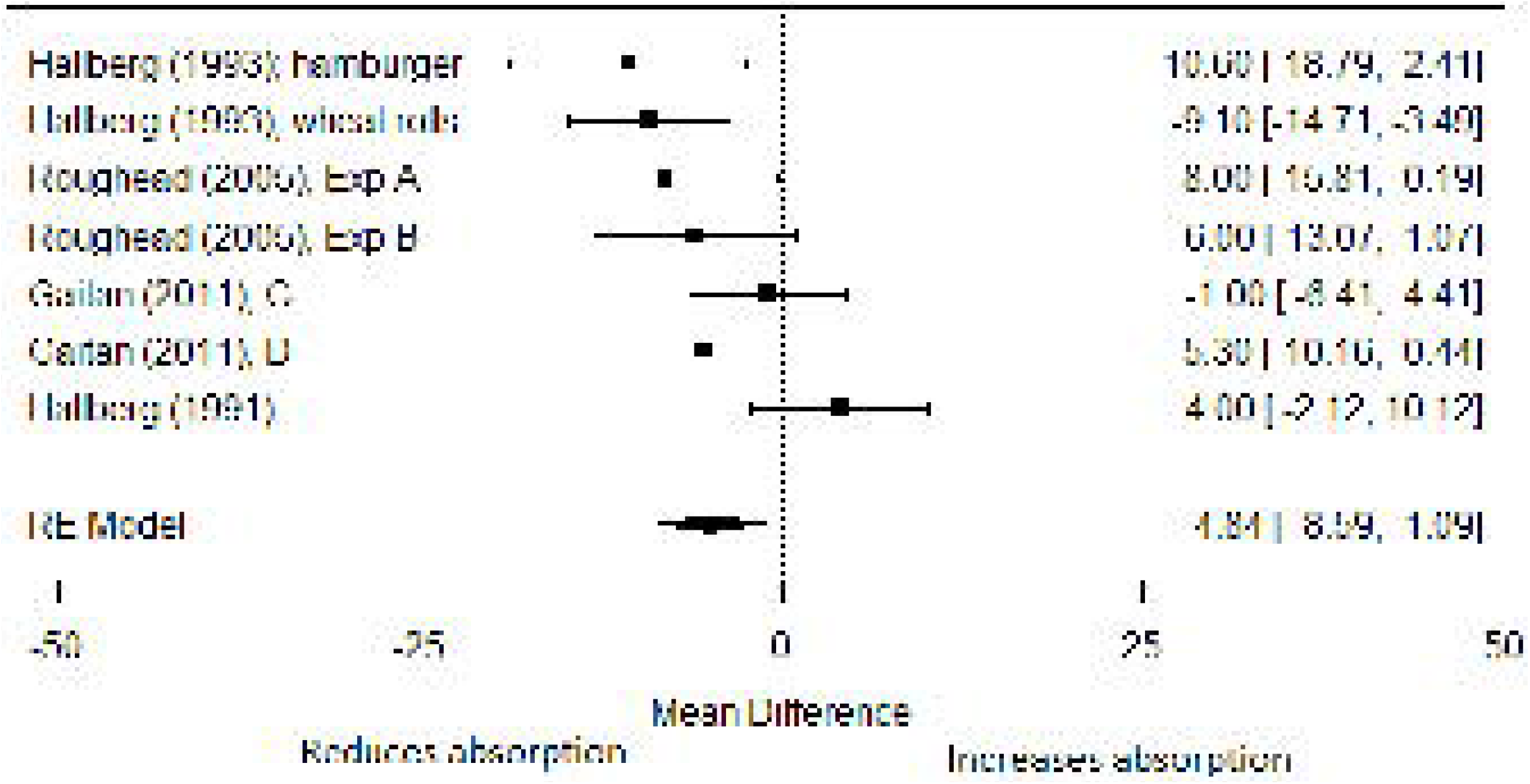
Forest plot for the relationship of calcium intake and heme iron absorption (highest vs least calcium intake)

We pooled 16 difference measures from 3 studies(1, 42, 48) using quadratic model-based meta-analysis to assess the presence of a non-linear dose response association. There was no evidence that the dose- response association of calcium intake on heme iron absorption was non-linear (*p-*value = 0.33). Heterogeneity was, however, substantial (I^2^=98%).

### Non-heme iron absorption

We pooled 11 estimates from 8 studies(2, 11, 22, 30, 34, 42, 43, 47, 48) to obtain weighted mean differences comparing the influence of high calcium intake on non-heme iron absorption relative to low calcium intake, and found that calcium intake was associated with a slightly lower non-heme iron absorption (**Figure 5**. WMD = -2.40, 95% CI: -4.98, 0.18; I^2^=98%, *p*-heterogeneity<0.0001), although the confidence limits included the null of 0. Follow-up of the included studies was from 1 – 15 days. Heterogeneity was not significantly explained by participant’s age (p-heterogeneity = 0.90), sex (p- heterogeneity=0.76), baseline serum ferritin (p-heterogeneity=0.73), nature of intervention (p- heterogeneity=0.43) or year of publication (p-heterogeneity=0.31). Visual inspection of the funnel plot suggested possible publication bias, though there was no evidence from Egger’s test of small study effects (Supplementary **Figure 4**, p-value=0.29).

**Figure 5.**
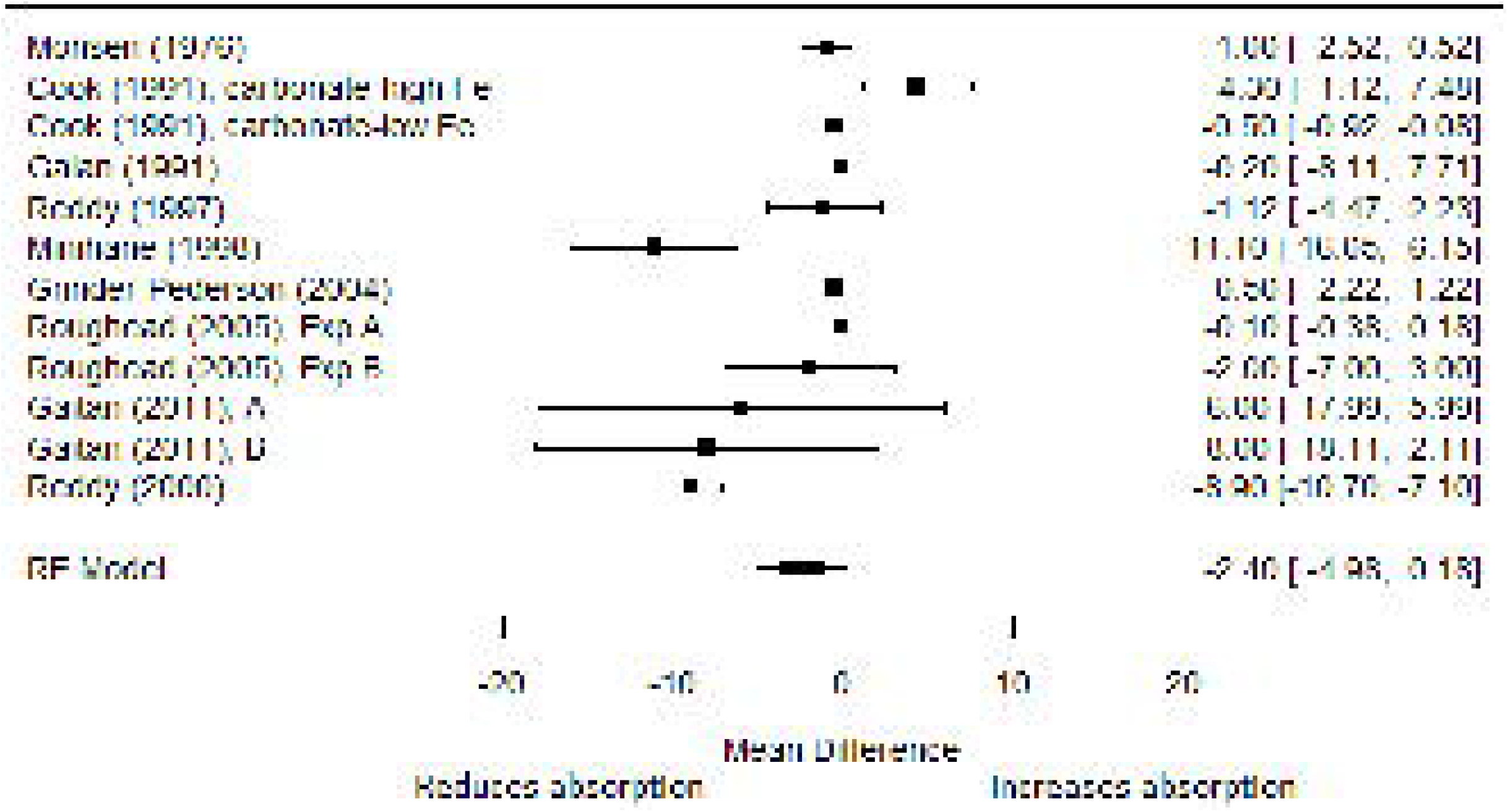
Forest plot for the relationship of calcium intake and non-heme iron absorption (highest vs least calcium intake)

We pooled 44 difference measures from 5 studies(22, 34, 42, 47, 48) using quadratic model-based meta- analysis. There was no evidence that the dose-response association of calcium intake on heme iron absorption was non-linear (*p-*value = 0.13). Heterogeneity was, however, substantial (I^2^=96%).

### Serum Ferritin

We pooled 13 estimates from 13 studies (10-14, 30-34, 50-52) to obtain weighted mean differences comparing the influence of high calcium intake and low calcium intake on serum ferritin concentration, and found no significant association of calcium intake and ferritin concentration (**Figure 6**. WMD = - 1.21µg/L, 95% CI: -5.47, 3.05). There was significant heterogeneity (I^2^=97%, *p*-heterogeneity<0.001).

**Figure 6.**
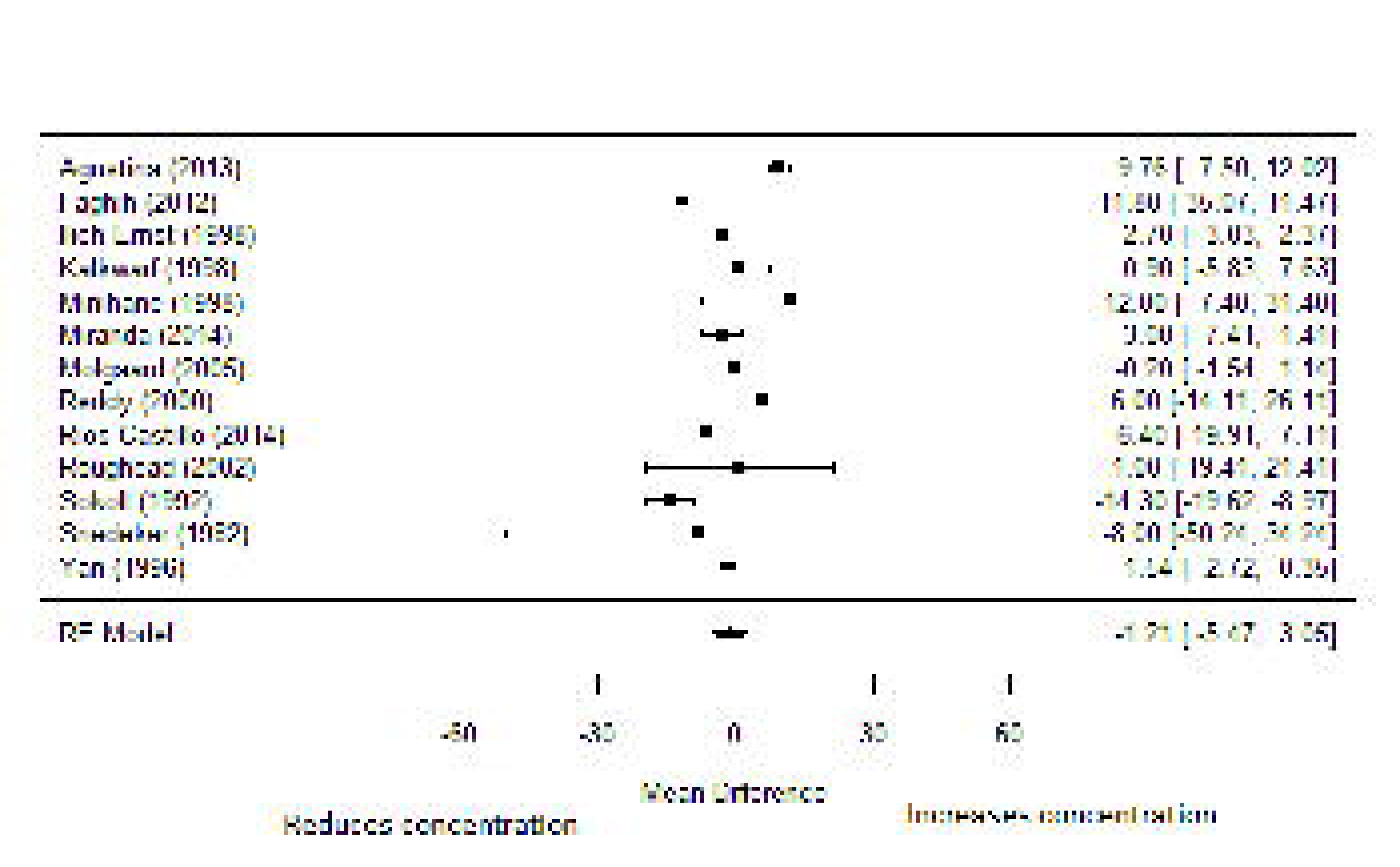
Forest plot for the relationship of calcium intake and serum ferritin (highest vs least calcium intake)

Heterogeneity was partly explained by whether the intervention included a calcium supplement or not (p-heterogeneity=0.005). Use of supplements was associated with 11.5 µg/L lower total iron absorption (95% CI: -19.5, -3.53) compared to calcium from diet alone. Heterogeneity was not significantly explained by participant’s age (p-heterogeneity=0.23), sex (p-heterogeneity=0.11), baseline serum ferritin (p-heterogeneity=0.26), hemoglobin concentration (p-heterogeneity=0.64), duration (p- heterogeneity=0.61) or year of study publication (p-heterogeneity=0.19). The duration of the interventions ranged from 2 – 52 weeks. Heterogeneity was not significantly explained by duration of follow-up (p-heterogeneity=0.19). There was no evidence from influence analysis that the pooled estimate was dominated by any of the individual studies. There was also no evidence of publication bias from visual inspection of funnel plots (**Supplementary Figure 5**) as well as Egger’s test for small study effects (p-value=0.92).

We pooled 32 difference measures from 3 short-term studies (12, 31, 47) using a quadratic model-based meta-analysis to explore dose-response relationship of calcium intake and iron absorption (**Figure 7)**. The dose of calcium in the included studies ranged from 22 – 1,250mg. The dose-response association of calcium intake with serum ferritin concentration was significant (*p-*value = 0.0004). Calcium intake was associated with reduced serum ferritin concentration as the dose increased. There was no meaningful heterogeneity (I^2^=0%).

**Figure 7.**
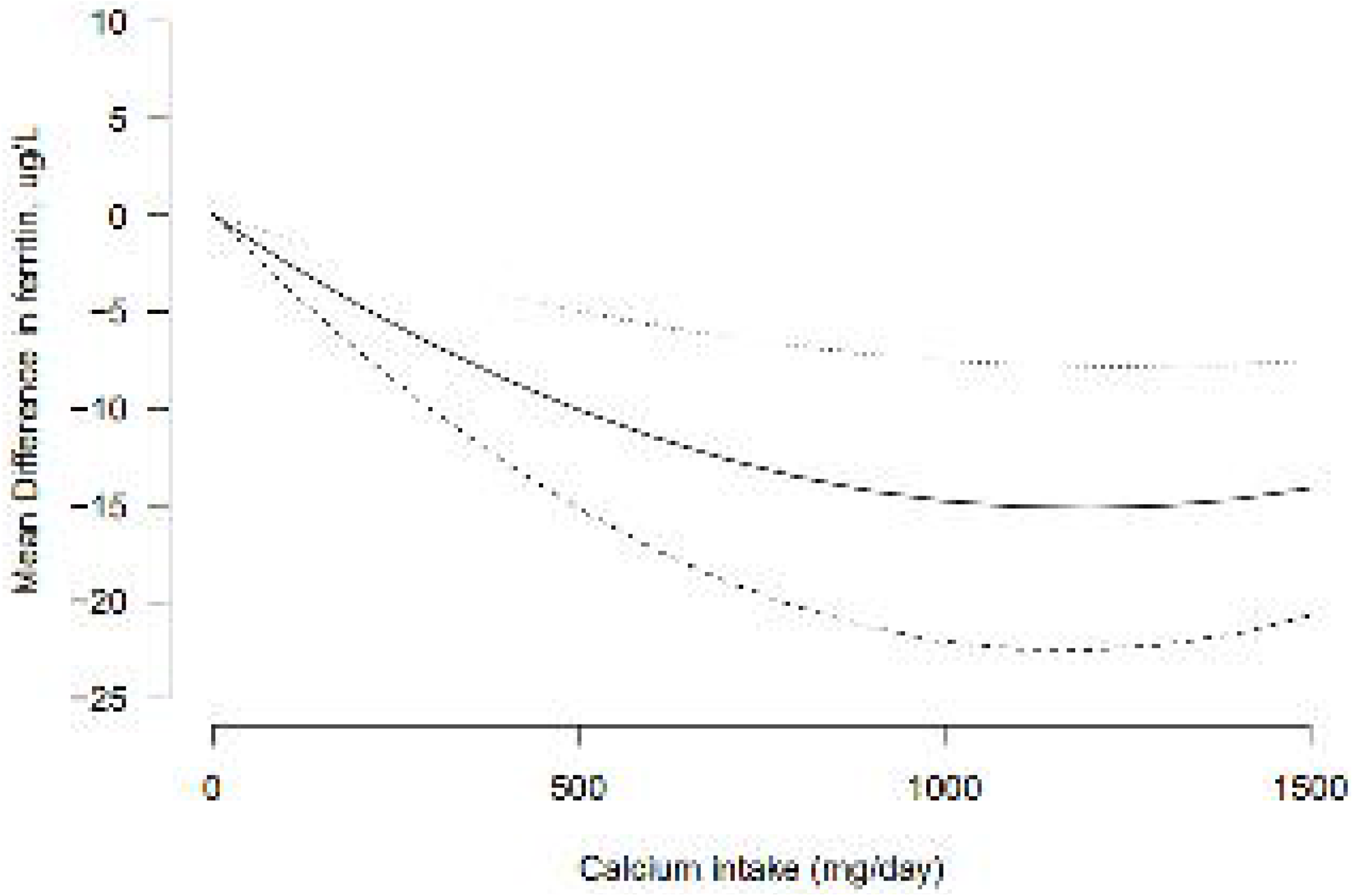
Dose-response relationship of calcium intake and serum ferritin concentration

### Hemoglobin concentration

We pooled 8 estimates from 6 studies (10, 12, 32, 33, 50, 51) to obtain weighted mean differences comparing the influence of high calcium intake on hemoglobin concentration relative to low calcium intake, and found that calcium intake no association with hemoglobin concentration (**Figure 8**. WMD = 0.44g/L, 95% CI: -0.90, 1.78). There was substantial heterogeneity (I^2^=77%, *p*-heterogeneity=0.001). The duration of intervention for the studies was 5 – 26wks, with one study that lasted 4 years. When we excluded the long-term study in sensitivity analysis, the WMD was 1.22 (95% CI: 0.37, 2.07).

**Figure 8.**
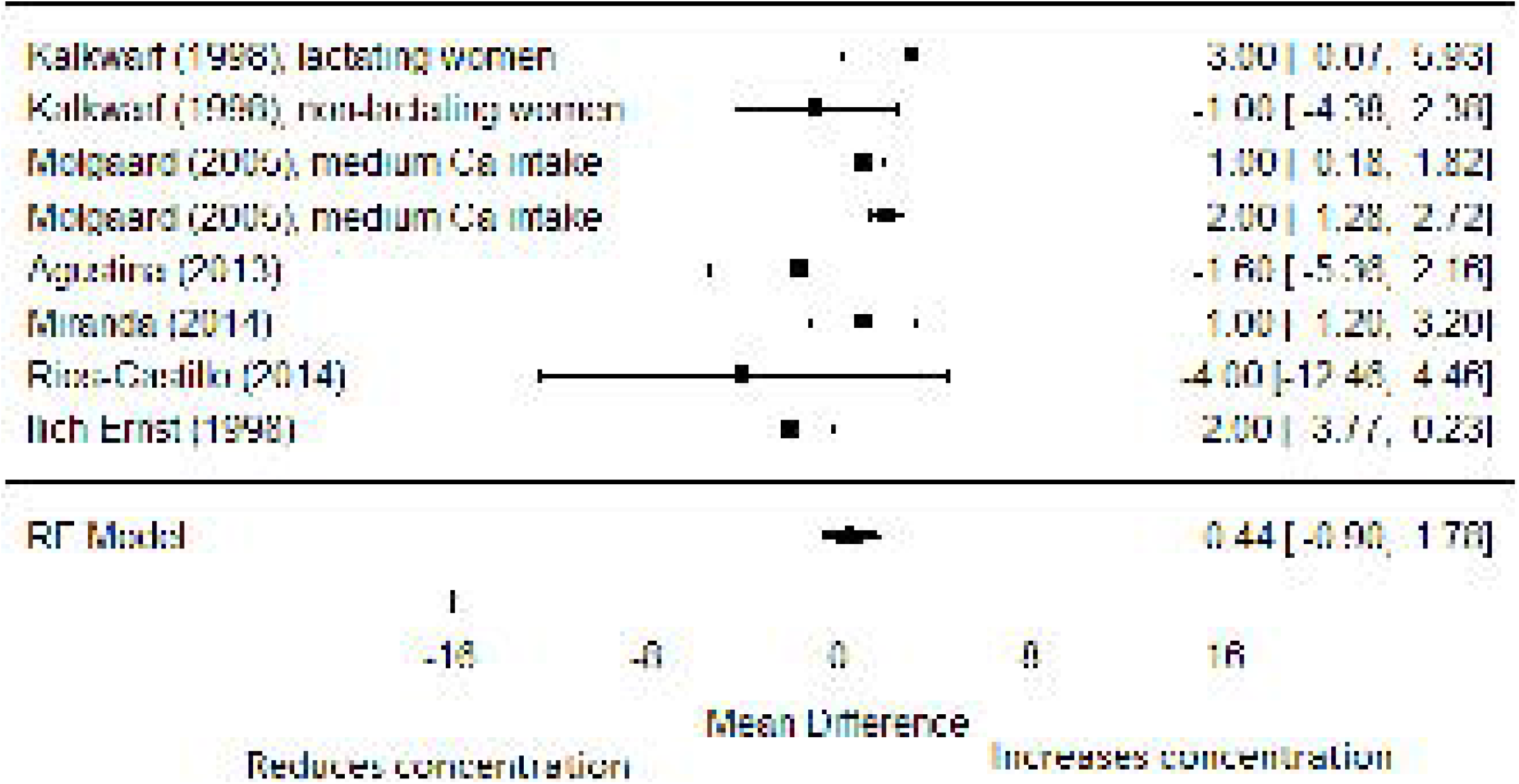
Forest plot for the relationship of calcium intake and hemoglobin concentration

Neither the funnel plot nor Egger’s test for small study effects (p=0.17) suggested possible publication bias (**Supplementary figure 6**). There were too few studies reporting multiple dose categories (n>2) and dose response analysis was not done.

### Sequencing of calcium intake

One study evaluated the extent to which the sequencing of calcium intake may influence iron absorption(45). Keeping daily dietary calcium intake constant, the investigators compared the iron absorption in 21 women who had dietary calcium at every meal to those who had no dietary calcium at lunch and dinner. They found that total, non-heme and heme iron absorption were greater in those who had no dietary calcium at lunch and dinner.

### Other outcomes

#### Iron retention

Three studies(3, 38, 41) examined the effect of calcium intake on iron retention. Among 11 boys and girls in Texas, USA who participated in a case-crossover study(38), there was no significant difference in the incorporation of iron into red blood cells between times when they were randomly assigned to high calcium diets compared to low calcium diets (mean difference: 1.0%, 95% CI: -6.2% to 4.2%).

Two crossover studies in Boston, USA studied the effect of calcium intake on iron absorption among postmenopausal women and found significantly lower retention of iron with intake of calcium from different sources. Among 13 women(3), intake of either calcium carbonate (percent reduction: 43.3% ±8.8, p=0.002) or hydroxyapatite (percent reduction: 45.9% ±10, p=0.003) led to significantly lower retention of iron, compared to placebo. Among 19 women(41), intake of milk (mean ±SD: 3.4 ±3.4) and calcium citrate-malate salt (mean ±SD: 6.0 ±4.2) led to significantly lower retention of iron (p <0.05), compared to placebo (mean ±SD: 8.3 ±4.6).

## Discussion

In this systematic review and meta-analysis, we summarized the human clinical studies that have examined the impact of calcium intake on iron status. We found that calcium intake was associated with overall reduced iron absorption and iron status in a dose dependent manner, but no impact on hemoglobin concentration in the long term. The effect estimates were small and unlikely to be biologically significant. Overall, the inverse relationship between calcium intake and iron status is unlikely to be clinically significant in free-living populations.

To the best of our knowledge, this is the first meta-analysis of the effect of calcium intake on iron status. The issue of calcium-iron interaction has assumed renewed significance in recent years, particularly in the context of low- and middle-income countries, due to recent WHO recommendations that calcium supplementation be included in routine ante-natal care for prevention of preeclampsia. The WHO currently recommends 1.5-2g of calcium supplements for pregnant women in communities with inadequate habitual calcium intake(53). The concern about the potential for a co-administered calcium and iron supplementation regimen to worsen the burden of anemia in pregnancy is also one of the justifications for calling for a lower dose for calcium supplementation(8, 54, 55), besides the issues of cost, logistical complexity and side effects.

Overall, our findings suggest that the magnitude of inhibition is unlikely to be clinically significant over time even if separated intake is not prescribed, but these findings should be interpreted and applied in the context of the weaknesses of the relevant body of work. We had set out to examine the impact of combined intake on hematological indices that have been robustly linked to functional consequences such as hemoglobin indices, over time periods that take iron homeostatic mechanisms into consideration. Unfortunately, the only study that compared combined intake to separated intake was a short-term study with iron absorption outcomes(45). There is a dearth of studies that have specifically examined the effects of concurrent ingestion of iron and calcium supplements compared to delayed intake of one or the other on biomarkers with well-established functional implications over extended periods.

This study has other notable limitations. The evidence base is mostly informed by studies that have examined the effect of different daily doses from meals and supplements on iron status over time, without indication of whether they were concurrently ingested or separated. In addition, the studies reviewed included subjects from a broader population and not pregnant women in low and middle- income countries only. In fact, most primary studies specifically excluded pregnant women; however the key population motivating the renewed debate about calcium-iron interaction is pregnant women. Moreover, the baseline iron status of the participants was not reported in many of the studies. It is well known that extant iron status is an important determinant of iron metabolism. Given the location of most of the studies in high income countries, it is plausible that the baseline iron status of participants might be higher than that of pregnant women in most communities in low- and middle-income countries. Furthermore, we examined hematologic status using serum ferritin and hemoglobin concentration. Too few studies have examined the effects of calcium intake on transferrin and other hematological markers and indices with important physiological and functional implications(12). The significant limitations of ferritin as a marker of iron status across populations is well known.

In conclusion, in the present systematic review and meta-analysis, we found statistically significant negative effect of calcium intake on iron status in the short term (≤ 90 days), but the magnitude of the effect was low and unlikely to be biologically significant, and longer-term studies consistently failed to find this effect. In fact, despite the negative effect of calcium intake on iron absorption, hemoglobin concentration was increased in this analysis. Our findings suggest that lowering the dose of calcium and iron supplement intake among pregnant women is unlikely to affect the anemia burden in the long-term. These findings should be interpreted with caution because of the significant heterogeneity and limitations of the underlying studies. There is a need for effectiveness trials comparing the effects of recommending separated supplement intake to combined intake among pregnant women in low and middle-income countries, with blood pressure and iron status as primary outcomes, and adherence to each supplement as intermediate outcomes.

## Data Availability

The datasheet for the meta-analysis are available upon request

## Abbreviations used in text

AIC: Akaike Information Criteria
CI: Confidence Interval
CINAHL: Current Nursing and Allied Health Literature
LMIC: Low- and middle-income countries
MeSH: Medical Subject Headings
PRISMA: Preferred Reporting Items for Systematic Reviews and Meta-analysis
RCT: Randomized controlled trial
RR: Relative risk
SD: Standard deviation
se: Standard error
WHO: World Health Organization
WMD: Weighted mean differences

## Acknowledgment

The authors are grateful to Dr Alessio Crippa of Karolinska Institutet for help with the R code for the dose-response analysis.

AIA and MOO designed research; All authors extracted data from primary studies; AIA and MOO analyzed data; AIA and MOO wrote the paper; MOO had primary responsibility for final content. All authors read and approved the final manuscript.

## Supplementary figures

Supplementary figure 1 – Funnel plot for the relationship of calcium intake and total iron absorption (highest vs least calcium intake)

Supplementary figure 2 – Funnel plot for the relationship of calcium intake and heme iron absorption (highest vs least calcium intake)

Supplementary figure 3 – Funnel plot for the relationship of calcium intake and non-heme iron absorption (highest vs least calcium intake)

Supplementary figure 4 – Funnel plot for the relationship of calcium intake and serum ferritin (highest vs least calcium intake)

Supplementary figure 5 – Funnel plot for the relationship of calcium intake and hemoglobin concentration

**Supplement 1.** PRISMA Checklist.

|  |  |  |  |
| --- | --- | --- | --- |
| **TITLE** |  |  |  |
| Title | 1 | Identify the report as a systematic review, meta-analysis, or both. | 1 |
| **ABSTRACT** |  |  |  |
| Structured summary | 2 | Provide a structured summary including, as applicable: background; objectives; data sources; study eligibility criteria, participants, and interventions; study appraisal and synthesis methods; results; limitations; conclusions and implications of key findings; systematic review registration number. | 2-3 |
| **INTRODUCTION** |  |  |  |
| Rationale | 3 | Describe the rationale for the review in the context of what is already known. | 5-6 |
| Objectives | 4 | Provide an explicit statement of questions being addressed with reference to participants, interventions, comparisons, outcomes, and study design (PICOS). | 6 |
| **METHODS** |  |  |  |
| Protocol and registration | 5 | Indicate if a review protocol exists, if and where it can be accessed (e.g., Web address), and, if available, provide registration information including registration number. | 7 |
| Eligibility criteria | 6 | Specify study characteristics (e.g., PICOS, length of follow-up) and report characteristics (e.g., years considered, language, publication status) used as criteria for eligibility, giving rationale. | 8-9 |
| Information sources | 7 | Describe all information sources (e.g., databases with dates of coverage, contact with study authors to identify additional studies) in the search and date last searched. | 7 |
| Search | 8 | Present full electronic search strategy for at least one database, including any limits used, such that it could be repeated. | 7 |
| Study selection | 9 | State the process for selecting studies (i.e., screening, eligibility, included in systematic review, and, if applicable, included in the meta-analysis). | 10 |
| Data collection process | 10 | Describe method of data extraction from reports (e.g., piloted forms, independently, in duplicate) and any processes for obtaining and confirming data from investigators. | 11 |
| Data items | 11 | List and define all variables for which data were sought (e.g., PICOS, funding sources) and any assumptions and simplifications made. | 11-12 |
| Risk of bias in individual studies | 12 | Describe methods used for assessing risk of bias of individual studies (including specification of whether this was done at the study or outcome level), and how this information is to be used in any data synthesis. | 14 |
| Summary measures | 13 | State the principal summary measures (e.g., risk ratio, difference in means). | 12 |
| Synthesis of results | 14 | Describe the methods of handling data and combining results of studies, if done, including measures of consistency (e.g., $I^2$) for each meta-analysis. | 13-14 |
| Risk of bias across studies | 15 | Specify any assessment of risk of bias that may affect the cumulative evidence (e.g., publication bias, selective reporting within studies). | 14 |
| Additional analyses | 16 | Describe methods of additional analyses (e.g., sensitivity or subgroup analyses, meta-regression), if done, indicating which were pre-specified. | 14 |
| **RESULTS** |  |  |  |
| Study selection | 17 | Give numbers of studies screened, assessed for eligibility, and included in the review, with reasons for exclusions at each stage, ideally with a flow diagram. | 15, 48, SI |
| Study characteristics | 18 | For each study, present characteristics for which data were extracted (e.g., study size, PICOS, follow-up period) and provide the citations. | 41-44 |
| Risk of bias within studies | 19 | Present data on risk of bias of each study and, if available, any outcome level assessment (see item 12). | 15-21 |
| Results of individual studies | 20 | For all outcomes considered (benefits or harms), present, for each study: (a) simple summary data for each intervention group (b) effect estimates and confidence intervals, ideally with a forest plot. | 15-21, SI, Figures |
| Synthesis of results | 21 | Present results of each meta-analysis done, including confidence intervals and measures of consistency. | 15-21 |
| Risk of bias across studies | 22 | Present results of any assessment of risk of bias across studies (see Item 15). | 15-21 |
| Additional analysis | 23 | Give results of additional analyses, if done (e.g., sensitivity or subgroup analyses, meta-regression [see Item 16]). | 15-21 |
| **DISCUSSION** |  |  |  |
| Summary of evidence | 24 | Summarize the main findings including the strength of evidence for each main outcome; consider their relevance to key groups (e.g., healthcare providers, users, and policy makers). | 22-23 |
| Limitations | 25 | Discuss limitations at study and outcome level (e.g., risk of bias), and at review-level (e.g., incomplete retrieval of identified research, reporting bias). | 24 |
| Conclusions | 26 | Provide a general interpretation of the results in the context of other evidence, and implications for future research. | 27-28 |
| **FUNDING** |  |  |  |
| Funding | 27 | Describe sources of funding for the systematic review and other support (e.g., supply of data); role of funders for the systematic review. | 29 |

**Supplement 2.** Search Strategy.

| Database | Search query | Original search | Updated search |
| --- | --- | --- | --- |
| PUBMED | ("Iron"[Mesh] AND "Calcium"[Mesh]) AND ("gastrointestinal absorption"[MeSH Terms] OR "biological availability"[Mesh] OR "inhibition"[All Fields] OR "interaction"[All Fields] OR "Iron-Binding Proteins"[Mesh] OR "Iron-Regulatory Proteins"[Mesh] OR "Hematologic Diseases"[Mesh]) | 505 | 47 |
| EMBASE | 'iron'/exp AND 'calcium'/exp AND ('bioavailability'/exp OR 'gastrointestinal absorption'/exp OR 'iron regulatory factor'/exp OR 'iron binding protein'/exp OR 'anemia'/exp) AND [embase]/lim NOT [medline]/lim NOT 'review'/it | 549 | 164 |

**Supplement 3.** Risk of Bias Assessment of randomized controlled trials.

| Author (year of publication) | Random sequence generation | Blinding of participants and personnel | Incomplete outcome data | Measurement of outcome | Selective reporting |
| --- | --- | --- | --- | --- | --- |
| Agustina (2013) | Low | High | High | Low | Low |
| Faghih (2012) | Low | High | High | Low | Low |
| Grinder-Pederson (2004) | Low | High | Low | Low | Low |
| Hallberg (1993) | Low | High | High | Low | Low |
| Israel Rios-Castillo (2014) | Low | Low | Low | Low | Low |
| Minihane (1998) | Low | High | Low | Low | Low |
| Miranda (2014) | Low | High | Low | Low | Low |
| Reddy (2000) | Some concerns | High | Low | Low | Low |
| Sokoll (1992) | Low | Low | Low | Low | Low |
| Walczyk (2014) | Low | High | Low | Low | Low |
| Yan (1996) | Low | High | Low | Low | Low |
| Mølgaard (2005) | Low | Low | Low | Low | Low |

**Supplement 4.**
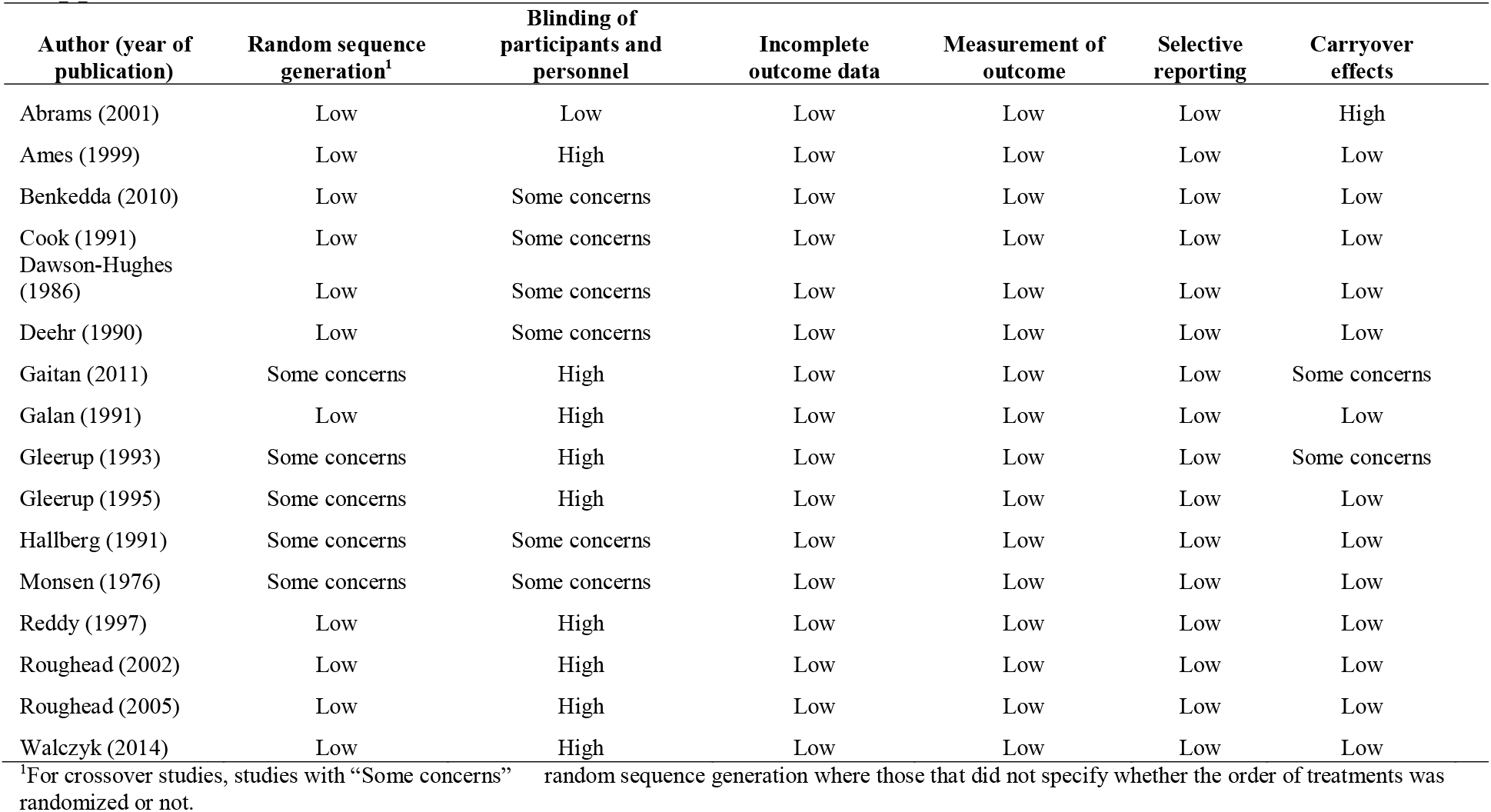
Risk of Bias Assessment for Crossover trials.

| Author (year of publication) | Random sequence generation<sup>1</sup> | Blinding of participants and personnel | Incomplete outcome data | Measurement of outcome | Selective reporting | Carryover effects |
| --- | --- | --- | --- | --- | --- | --- |
| Abrams (2001) | Low | Low | Low | Low | Low | High |
| Ames (1999) | Low | High | Low | Low | Low | Low |
| Benkedda (2010) | Low | Some concerns | Low | Low | Low | Low |
| Cook (1991) | Low | Some concerns | Low | Low | Low | Low |
| Dawson-Hughes (1986) | Low | Some concerns | Low | Low | Low | Low |
| Deehr (1990) | Low | Some concerns | Low | Low | Low | Low |
| Gaitan (2011) | Some concerns | High | Low | Low | Low | Some concerns |
| Galan (1991) | Low | High | Low | Low | Low | Low |
| Gleerup (1993) | Some concerns | High | Low | Low | Low | Some concerns |
| Gleerup (1995) | Some concerns | High | Low | Low | Low | Low |
| Hallberg (1991) | Some concerns | Some concerns | Low | Low | Low | Low |
| Monsen (1976) | Some concerns | Some concerns | Low | Low | Low | Low |
| Reddy (1997) | Low | High | Low | Low | Low | Low |
| Roughead (2002) | Low | High | Low | Low | Low | Low |
| Roughead (2005) | Low | High | Low | Low | Low | Low |
| Walczyk (2014) | Low | High | Low | Low | Low | Low |

<sup>1</sup>For crossover studies, studies with “Some concerns” random sequence generation where those that did not specify whether the order of treatments was randomized or not.

## Notes

### Competing Interest Statement

The authors have declared no competing interest.

### Funding Statement

No funding was received for this work at all.

### Author Declarations

This is a meta-analysis of publicly available studies, and would not warrant an IRB review or exemption.

